# Impact of marginal donor to marginal recipient kidney transplant on delayed graft function and outcome

**DOI:** 10.1101/2023.03.27.23287806

**Authors:** Federica Bocchi, Guido Beldi, Christian Kuhn, Federico Storni, Nathalie Müller, Daniel Sidler

## Abstract

The demographics of donor and recipient candidates for kidney transplantation (KT) have substantially changed. Recipients tend to be older and polymorbid and KT to marginal recipients is associated with delayed graft function (DGF), prolonged hospitalization, inferior long-term allograft function, and poorer patient survival. In parallel, donors are also older, suffer from several comorbidities, and donations coming from circulatory death (DCD) predominate, which in turn leads to early and late complications. However, it is unclear how donor and recipient risk factors interact. In this retrospective cohort study, we assess the overall and combined impact of a KT from marginal donors to marginal recipients. We focused on: 1) DGF; 2) hospital stay and number of dialysis days after KT and 3) allograft function at 6 months. Among the 369 KT included, the overall DGF rate was 25% (n = 92) and median time from reperfusion to DGF resolution was 7.8 days (IQR: 3.0-13.8 days). Overall, patients received four dialysis sessions (IQR: 2-8). The combination of pre-KT anuria (< 200 ml/24h, 32%) and DCD procurement (14%) was significantly associated with DGF, length of hospital stay, and severe perioperative complications, predominantly in recipients 50 years and older.

## INTRODUCTION

The prevalence of end-stage kidney disease (ESKD) has increased substantially over the last years (1). Kidney transplantation (KT) is the preferred renal replacement therapy for eligible patients. It improves survival, reduces morbidity and provides a quality of life benefit when compared to hemodialysis (2), (3), (4). However, due to the burden of ESKD, the shortage of living and deceased donor candidates, and stringent eligibility criteria, the availability of suitable organs is still the limiting factor for KT (5).

In KT, allografts are transferred from the donor to the recipient with minimal time delay to reduce reperfusion injury and subsequent microvascular damage, thereby preserving graft viability. Delayed graft function (DGF), a major clinical challenge, is mainly defined on serum creatinine values or the requirement of renal replacement therapy within the first week after KT. It’s incidence reported worldwide varies between transplant centers, ranging approximately from 4-10% in living donors KT to 19-70% in deceased KT, also reflecting the heterogeneity of its definition (6). DGF has a negative connotation because it is associated with early acute rejection, peri-operative mortality, prolonged hospitalization, increased cost of care, and shortened allograft survival (7).

Over the last years, the characteristics of recipient and donor candidates for KT have changed substantially. Recipients tend to be older and polymorbid, often having spent several years on dialysis, and frequently are without residual urine production. KT to marginal recipients is associated with DGF, prolonged hospitalization, inferior long-term allograft function, and poorer patient survival (8), (9).

In parallel, donors nowadays are also older, suffer from several comorbidities (especially cardiovascular disease) and donation after cardiac death (DCD) predominate, which in turn leads to early and late complications (10), (11), (12).

For these reasons, the incidence of DGF is expected to increase in the coming years, making the identification of potential risk factors an interesting prevention tool. Several risk factors for DGF (cold ischemia time, donor and recipient age, cause of donor death) have already been identified (6). Furthermore, several attempts have been made to predict the risk of DGF, such as nomograms based on donor and recipient risk factors (13), preimplantation biopsies (with (14) and without (15) transcriptomics), or urinary biomarkers measured early after transplantation (16) (17). However, these methods may not be very accurate, might show a high interobserver variability (preimplantation biopsies) or are not readily available at time of allocation (transcriptomics, urinary biomarkers). A better understanding of variables influencing DGF that are already known at time of allocation would allow better use of resource, improve the KT outcomes, and thus promote widespread utilization of kidney at risk for DGF.

In this study, we employed data from a single-center comprehensive KT cohort to assess the overall and combined impact of KT from marginal donors to marginal recipients. We focused on 1) DGF; 2) hospital stay duration and number of dialysis days after KT; 3) allograft function at 6 months; and 4) patient and allograft survival.

## MATERIALS AND METHODS

### Study design and population

This study is an analysis of prospective registry data. It covers the period from January 5th 2008 to April 7th 2022 and includes all patients (n = 627) who received a KT at the university hospital of Bern, Switzerland. Median follow-up time was 5.41 years (IQR: 2.06-8.31 years). Recipient and donor characteristics were collected from the clinical information platform and laboratory analyses extracted via Insel Data Science Center (IDSC). Patient data of deceased donors were extracted from the Swiss Organ Allocation System (SOAS). Pre- and post-KT dialysis sessions parameters in recipients were extracted from the dialysis therapy management database.

### Delayed graft function

DGF was defined as the need for at least one dialysis session within one week after KT (time between reperfusion and first dialysis start), regardless of indication (volume overload, hyperkalemia, azotemia). Time from reperfusion to first dialysis and to end of last dialysis was calculated, as well as number of dialysis sessions, cumulative dialysis time and cumulative ultrafiltration.

### Endpoints and co-variates

Anuria was defined as residual urine production of less than 200 ml/24h at the time of KT. Following co-variables were also considered: donor characteristics (age, acute kidney injury, resuscitation, cardiovascular comorbidities, type of organ procurement (donation after cardiac or brain death, DCD or DBD), use of hypothermic machine perfusion), recipient characteristics (age, dialysis history, residual urine volume, previous KT history), cold and warm ischemia time, and first warm ischemia in the case of DCD. First warm ischemia was defined as 0 min in case of DBD procurement. Finally, the type of induction therapy (depleting vs. non-depleting) was assessed. In-house complications were assessed according to the Clavien-Dindo classification (18). Post-operative biopsies of the kidney allografts and dialyses were excluded as relevant Clavien-Dindo complications for all patients in this study.

The primary outcome of the study was the risk for the dichotomous event of DGF after KT. Secondary outcomes were the risk for major complications, length of hospital stay, estimated glomerular filtration rate (eGFR) at indicated time points, and time to graft loss or patient death. eGFR, given in ml/min/1.73 m^2^ was calculated using the Chronic Kidney Disease Epidemiology Collaboration 2009 (CKD-EPI) creatinine equation (19) at 6 months after KT. For patients with graft loss before 6 month, eGFR was set to at 0 ml/min/1.73m^2^. Further endpoints included the association of pre-KT dialysis (within 12 hours before KT), depleting induction therapy, use of hypothermic machine perfusion, and cold ischemia time with DGF events.

### Exclusion criteria

Participants were excluded from the analysis if (1) they had a living donor transplantation (LDT); (2) were multi-organ recipient; (3) were paediatric recipients (age < 18 years old); or (4) had a primary graft non-function (absence of dialysis-independent graft function until terminal loss).

### Patient and public involvement

Neither patients nor the public were involved in the design, conduct, or analysis of this study.

### Statistical analysis

Results were reported as number of participants (percentage) for categorical data and median (interquartile range) for continuous data.

The odds ratio (OR) for a DGF event was calculated from a full model of donor-, recipient- and procedural-derived risk factors (donor: age, resuscitation status, DCD procurement [yes, no], comorbidities [any of hypertension, diabetes mellitus or cardiovascular disease]; recipient: age, dialysis vintage [years], anuria [yes, no]; procedural: cold and warm ischemia time). Significant factors were retested in a reduced model. In a second approach, log-transformed first warm ischemia time (per 15 min) and residual urine volume (per 100 ml/24h) were considered as quantitative parameters for model calculations. From these results, the risk for DGF was refitted for the KT cohort using the following parameters: donor and recipient age, DCD parameters (qualitative and quantitative), residual urine volume (qualitative and quantitative).

The discriminative powers of the reduced models were calculated by the area under the receiver operating characteristic curve (AUC-ROC) to predict DGF events. The models were further internally validated by calibration analysis and respective Brier scores calculated. The Brier score is the mean squared difference between the predicted probability and the actual outcome with a lower Brier score (scale: 0-1) reflecting a better calibrated model.

Time-to-event analyses were performed including length of hospital stay, time to first re-hospitalization, time to graft loss, patient death or the composite endpoint of patient death or graft loss, and plotted as Kaplan-Meier curves and cumulative incidence curves, respectively. The incidence of perioperative complications (Clavien-Dindo grade IIIb or higher) was calculated for the respective groups, and further sub-analyzed for recipients younger or older than 50 years at the time of KT. Differences were measured using chi-square tests.

Kidney function 6 months after KT was illustrated among the groups, and differences compared using Mann-Whitney U tests.

The association between DGF events and pre-KT interventions (dialysis, hypothermic machine perfusion, induction therapy, cold ischemia time) was analysed by logistic regression adjusted for residual urine volume and DCD procurement as important cofounders.

Results were expressed as multivariable-adjusted mean±SD for categorical values and as adjusted regression coefficient for continuous variables. A two-tailed p<0.05 was considered statistically significant.

Statistical analyses were performed using R (version 4.0.3) and R Studio (version 1.3.1093).

## RESULTS

### Selection procedure and overall characteristics of participants

Of the initial 627 participants, 369 (58.8%) were included. The selection procedure is summarised in supplementary figure 1. The majority of participants were male, and donor and recipient age were 57 years (IQR: 44-67) and 58 years (IQR: 49-66), respectively. Among donors, arterial hypertension was present in 31% of cases, 14% of KT came from DCD procurement, and the use of a hypothermic machine perfusion was reported only in few cases (14%). Regarding the recipients, only a minority had a preemptive KT (8.5%), and the rest had already spent an average of 3.24 years (IQR 2.08-4.44) on dialysis, with 32% of participants already being anuric. Warm and cold ischemia times were 29 minutes (IQR: 24-34) and 8.12 hours (IQR: 6.40-10.40), respectively (table 1a and 1b).

**Table 1a.** Baseline donor characteristics.

|  | **Donors** |
| --- | --- |
|  | **N = 334** |
| **Age (years)** | 57 (44, 67) |
| **Gender** |  |
| Male | 180 (54%) |
| **Procurement** |  |
| DCD (yes) | 47 (14%) |
| KDPI (%) | 85 (60, 98) |
| Resuscitation (yes) | 104 (31%) |
| AKI (yes) | 109 (34%) |
| CVD (yes) | 70 (21%) |
| Hypertension (yes) | 103 (31%) |
| Diabetes mellitus (yes) | 25 (7.6%) |
| Hypothermic machine perfusion (yes) | 45 (14%) |

DCD: Donation after Circulatory Death; KDPI: Kidney Donor Profile Index; AKI: Acute Kidney Injury. CVD: Cardiovascular Disease.

**Table 1b.** Baseline recipient characteristics.

|  | **Recipients** |
| --- | --- |
|  | **N = 392** |
| **Age (years)** | 58 (49, 66) |
| **Gender** |  |
| Male | 232 (63%) |
| **Renal replacement therapy** |  |
| Time (years) | 3.24 (2.08, 4.44) |
| Residual urine (ml/24h) | 800 (0, 1.800) |
| Anuria (< 200 ml/24h) | 111 (32%) |
| Previous KT | 71 (19%) |
| **Transplantation** |  |
| Dialysis before KT (yes) | 118 (32%) |
| Preemptive KT (yes) | 29 (8.5%) |
| Cold ischemia time (hours) | 8.12 (6.40, 10.40) |
| Warm ischemia time (min) | 29 (24, 34) |
| First warm ischemia time (min) [in DCD] | 27 (22, 34) |
| Hypothermic machine perfusion (yes) | 51 (14%) |
| **Induction** |  |
| Depleting induction therapy (yes) | 28 (18%) |

KT: Kidney Transplantation; DCD: Donation after Circulatory Death.

### Delayed graft function and risk factors

The overall frequency of DGF was 25% (n = 92). The absolute and relative frequencies of DGF between 2008 and 2022 are illustrated in figure 1a and b. Median time from reperfusion to end of last dialysis was 7.8 days (IQR: 3.0-13.8) (figure 2c). Overall, patients required a median of 4 dialyses (IQR: 2-8) with a median total dialysis time per patient of 12.4 hours (IQR 5.7-26.6). More details are provided in table 2. In a regression model, we identified DCD procurement alongside anuria as relevant risk factors for DGF. These factors remained statistically significant when analyzed in a reduced model including donor and recipient age, and independently predicted DGF events with an AUC of 70% (CI: 64-77%, p<0.05). Our model predicted DGF events from 15% in DBD KT for recipients with intact diuresis to 71% in DCD KT to anuric recipients (table 3, figure 2a-b). Calibration of observed and estimated DGF probabilities revealed that prediction was reliable over a wide range with a Brier Score of 0.16. The model overestimated the risk of DGF in patients with an intermediate predicted likelihood (figure 2c). Similarly, quantitative values of first warm ischemia time and residual urine volume were significantly associated with DGF and gradually predicted events with an AUC of 69% (CI: 63-76%) with an analogous calibration performance (figure 2d-f).

**Figure 1:**
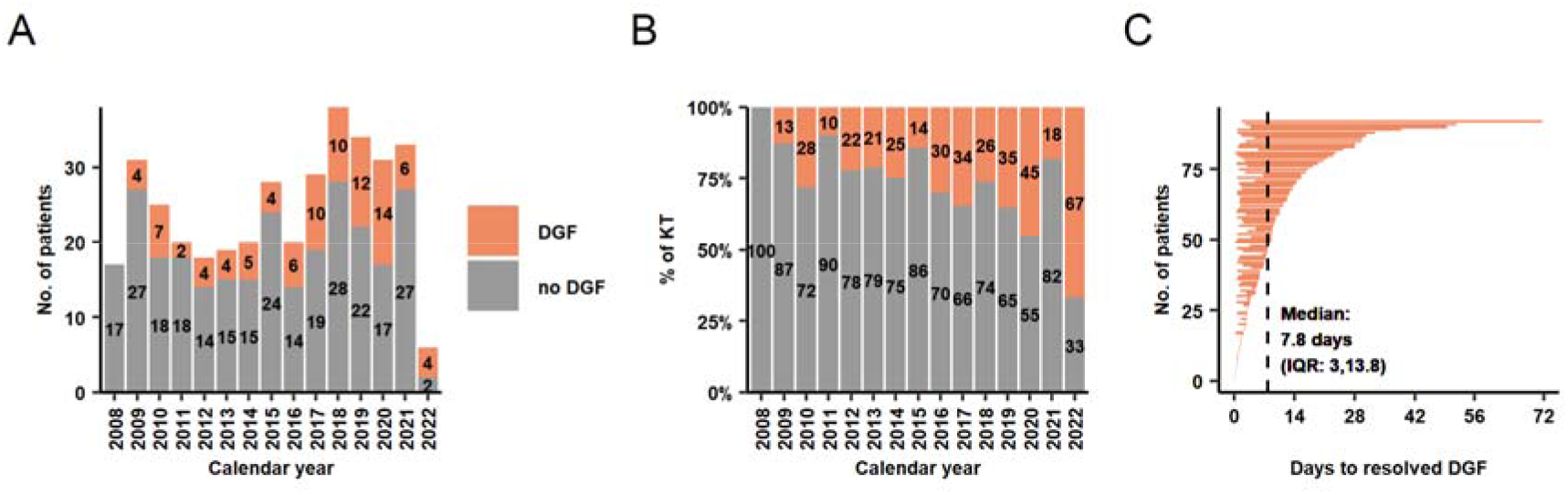
a-b) Absolute and relative frequency of KT with and without DGF during the study period. c) Time on dialysis after reperfusion for 92 patients with DGF. Median time from reperfusion to end of last dialysis was 7.8 days. DGF: Delayed Graft Function; KT: Kidney Transplantation.

**Figure 2.**
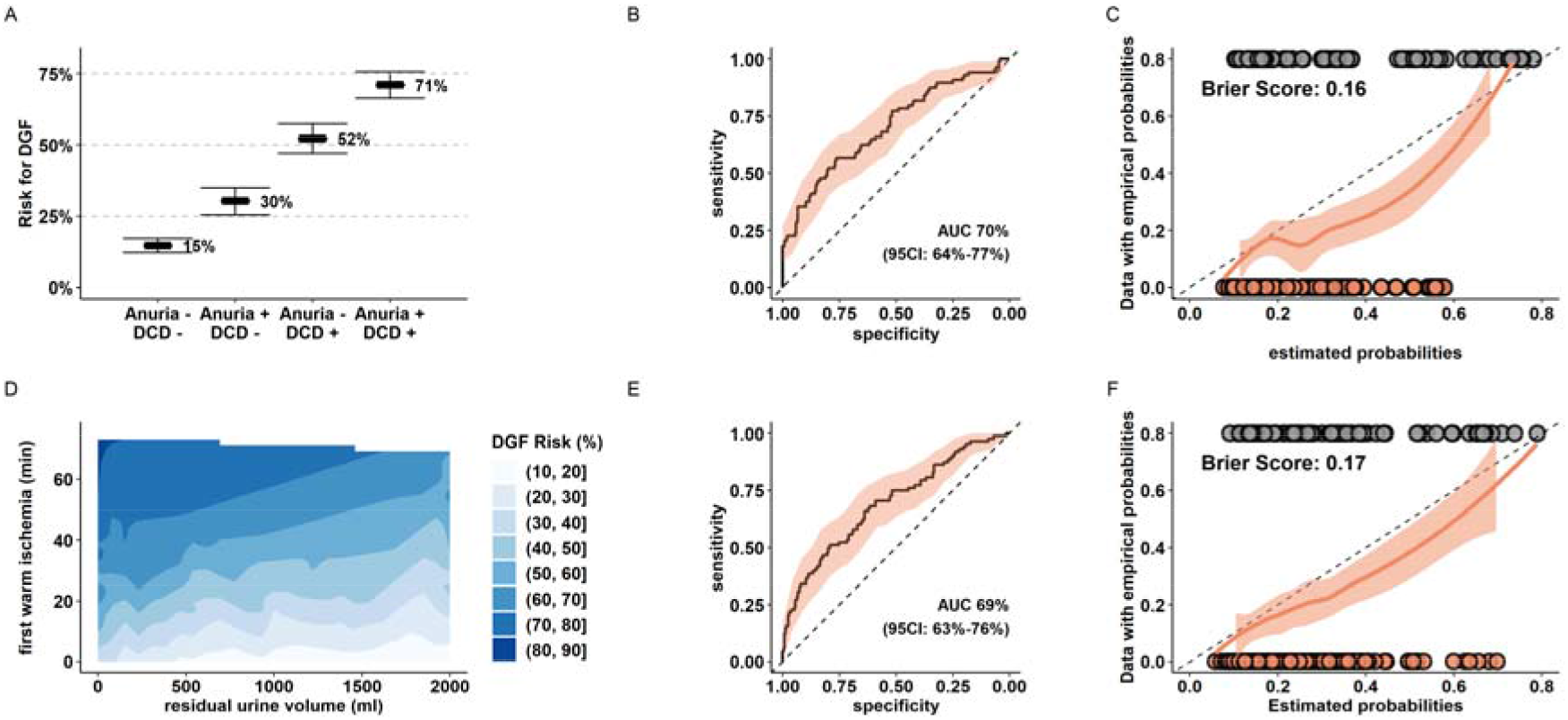
Prediction models for DGF in the Bernese cohort. a-c) Prediction model for DGF with organ procurement (DBD versus DCD), residual urine volume (≥ 200 ml/24h versus < 200 ml/24h), donor and recipient age as exploratory parameters. d-f) Prediction model for DGF with first warm ischemia time (per 15 min), residual urine volume (per 100 ml), donor and recipient age as quantitative exploratory variables. b, e) ROC analysis and calculated AUC for the respective models. c, f) Calibration analysis of empirical, estimated probabilities and Brier scores from the reduced models applied to the entire cohort. d) Estimated risk for DGF based on the reduced model. DGF: Delayed Graft Function, DCD: Donation after Circulatory Death; DBD: Donation after Brain Death.

**Table 2.** Characteristics of KT recipient with DGF. KT: Kidney Transplantation; DGF: Delayed Graft Function.

|  | **KT recipients with DGF** |
| --- | --- |
|  | **N = 92** |
| Total post-KT dialysis sessions (no) | 4 (2, 8) |
| Total time on dialysis (hours) | 12.4 (5.7, 26.6) |
| Average ultrafiltration volume on dialysis (liters) | 2.51 (1.71, 3.21) |
| At least one dialysis with ultrafiltration | 87 (95%) |
| Time to first dialysis (days) | 1.3 (0.8, 2.5) |
| Time to last dialysis (days) | 7.98 (3.0, 13.8) |

KT: Kidney Transplantation; DGF: Delayed Graft Function.

**Table 3.** Logistic regression model for DGF (yes/no) as dependent variable and donor-, recipient- and procedural factors as independent variables (full model). Donor and recipient age, as well as significant variables (DCD procurement, residual urine volume < 200 ml/24h) from the full model were used for a subsequent reduced model. DGF: Delayed Graft Function; DCD: Donation after Circulatory Death.

|  |  | Full model |  |  | Reduced model |  |  |
| --- | --- | --- | --- | --- | --- | --- | --- |
|  |  | OR | 95% CI | p-value | OR | 95% CI | p-value |
| Recipient | Age (per decade) | 1.18 | 0.92, 1.52 | 0.2 | 1.16 | 0.93, 1.46 | 0.2 |
|  | Dialysis time (per year) | 1.03 | 0.91, 1.17 | 0.6 | - | - | - |
|  | Anuria | 2.33 | 1.26, 4.34 | **0.007** | 2.67 | 1.54, 4.67 | **< 0.001** |
| Donor | Age (per decade) | 1.09 | 0.89, 1.33 | 0.4 | 1.04 | 0.88, 1.23 | 0.7 |
|  | AKI (yes) | 1.74 | 0.95, 3.20 | 0.074 | - | - | - |
|  | Resuscitation (yes) | 1.27 | 0.68, 2.37 | 0.4 | - | - | - |
|  | DCD (yes) | 7.25 | 3.55, 15.3 | **< 0.001** | 6.08 | 3.25, 11.6 | **< 0.001** |
|  | Comorbidities (yes) | 1.12 | 0.61, 2.09 | 0.7 | - | - | - |
|  | Cold ischemia time (per hour) | 1.04 | 0.95, 1.13 | 0.4 | - | - | - |
|  | Warm ischemia time (per 15 min) | 1.33 | 0.80, 2.20 | 0.3 | - | - | - |

AKI: Acute Kidney Injury; DCD: Donation after Circulatory Death. Comorbidities: any of hypertension, diabetes mellitus, cardiovascular disease.

### Complications, hospitalization duration and kidney function outcome

Severe perioperative complications (Clavien-Dingi grade IIIb or higher) were increased up to fivefold (4% in DBD KT to recipients without anuria versus 20% in DCD KT to recipients with anuria). Strikingly, this ratio further increased in a subgroup of recipients aged 50 years and older at time of KT with an incidence of 33% in the highest risk group (figure 3a-b). The majority of such complications was medical (11/23, 48%, mostly vascular events and respiratory failure), followed by surgical (8/23, 35%, mostly revisions and gastrointestinal interventions).

**Figure 3.**
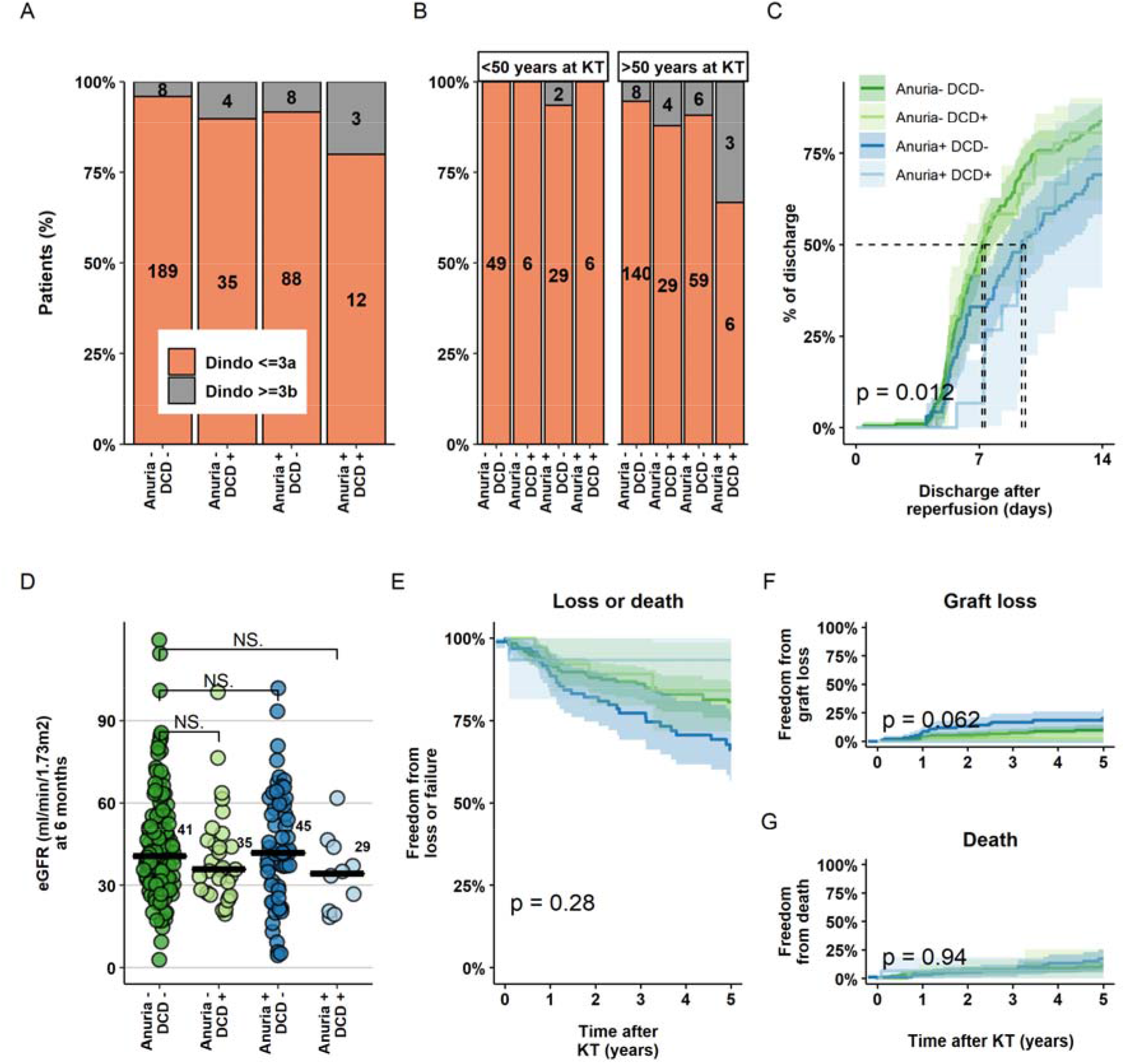
Perioperative complications, hospital stay duration and eGFR at 6 months after KT. a) Absolute and relative frequencies of Clavien-Dindo complications ≥ IIIb (interventions under complete anaesthesia, intensive care unit treatment, death) in the various groups. b) Absolute and relative rate of Clavien-Dindo grade ≥ IIIb complications stratified for recipients younger and older than 50 year at time of KT. c) Duration until first hospital discharge after KT for the various groups, d-e) Kidney function outcome. KT: Kidney Transplantation, eGFR: estimated Glomerular Filtration Rate, DCD: Donation after Circulatory Death.

Median hospital duration was overall short, but significantly longer in anuric recipients of both DBD and DCD KT (non anuric recipients: 7.17-7.33 days, anuric recipients: 9.42-9.62 days, p<0.05) (figure 3c). Finally, KT function at 6 months and long-term patient and graft survival were comparable among the risk groups (figure 3d-e).

Overall, hypothermic machine perfusion, depleting induction therapy, and dialysis treatment within 12 hours before KT were infrequent and ranged from 15- to 34% of all KT among the groups. In a retrospective analysis and after correction for DCD procurement and residual urine volume, none of these interventions reduced DGF probability (figure 4, supplementary table 1).

**Figure 4.**
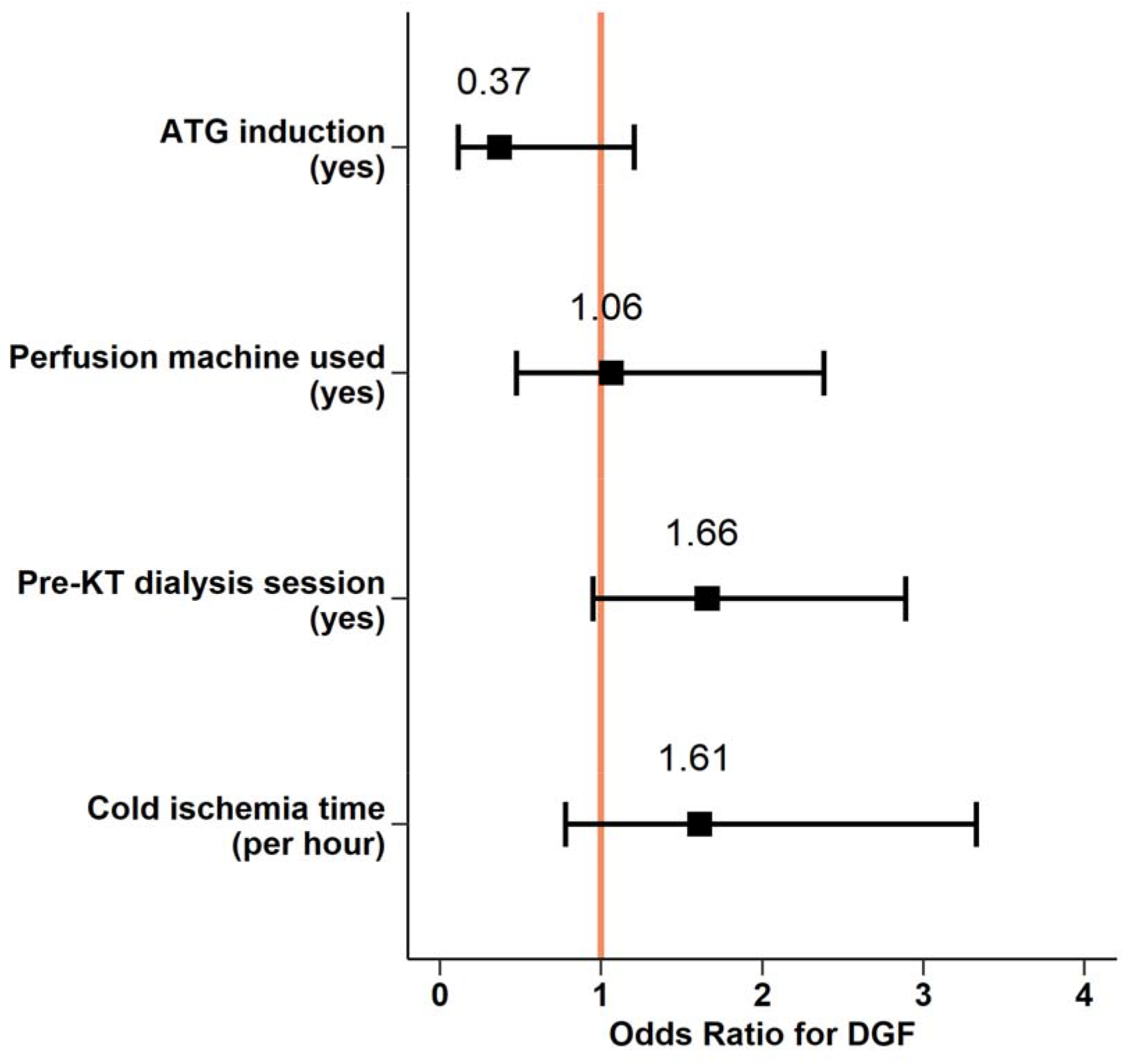
Perioperative interventions and DGF risk. Logistic regression model predicting DGF events from exploratory given variables and log-transformed cold ischemia time and residual urine volume as co-founding parameters. DGF: Delayed Graft Function; KT: Kidney Transplantation; ATG: Antithymocyte Globulin.

## DISCUSSION

To our knowledge, this is the first study assessing the incidence, characteristics and outcome of DGF in a Swiss population. In a study sample of 369 participants, we found that DCD procurement and residual urine volume were strongly associated with DGF.

Considering the ESKD burden, the severe shortage of high-quality organs for KT and the use of nonstandard kidney from expanded criteria donors, the incidence of DGF will remain high in the next years. For this reason, preventive measures in order to prevent DGF are under consideration.

DGF was a rather frequent event and occurred in 25% of patients receiving a KT. Similar results were recently published by Jadlowiec et al (20). As described in a previous cohort (21), we identified DCD procurement as an important predictor of DGF. This procurement approach is linked to a variable first warm ischemia time, during which relevant hypoxic damage occurs in the kidney with subsequent reperfusion injury and development of acute tubular necrosis. To prevent DGF, one of the best-known preventive measures is to decrease warm and cold ischemia time. If on the one hand surgeons have a concrete way to act and reduce the risk of DGF on the other hand also hypothermic preservation machines provide an additional benefit. Several studies have proved the superiority of hypothermic machine perfusion over standard cold storage in terms of DGF (22), (23) (24), yet its impact on long-term outcome is less clear. In our study, only a minority of KT underwent a hypothermic machine perfusion, and this intervention was not independently associated with a relevant reduction of DGF events (OR 1.06, CI: 0.46-2.33, p = 0.9).

The duration of kidney replacement therapy before KT is an important risk factor associated with DGF (25) (26). It is not fully understood whether the time spent on dialysis alters the recipient’s environment such that the incidence of ischemia-reperfusion injury increases and consequently influences the risk of DGF, or whether this is directly related to the progressive loss of residual urine volume. We found that the recipient’s residual urine volume, rather than the dialysis history, was significantly associated with the risk of DGF. This is intuitive, since decreasing residual diuresis is associated with minimal residual kidney function and, more important, with consequently poor volume control. Interestingly, although other factors (including cold- and warm ischemia time, donor and recipient age, or comorbidities) were (insignificantly) associated with DGF risk in our study, their influence was much smaller compared to DCD procurement and residual urine volume.

We demonstrate overall stable and sufficient graft function in marginal-to-marginal KT, although 6-month eGFR tends to be lower and is likely associated with accelerated graft deterioration. We further show that marginal-to-marginal KT is associated with longer primary hospitalization duration, which compromises rehabilitation and significantly increases the burden on the health care system. Finally, we found that early complications are uncommon but increased yet some up to fivefold in KT settings from DCD donors to anuric recipients. This risk is notably highest in recipients aged 50 years and older. This is important since perioperative complications after KT are associated with significant mid- and long-term morbidity. As reported by Jadlowiec et al., the readmission rate as well as the complications in this group of patients may explain the less favorable outcomes at 6-months and the lower graft survival (20). This would suggest that others factors, independent of DGF, could have a considerable impact on KT outcomes.

Identifying factors contributing to KT outcome is essential. For example, minimizing perioperative stress on the recipient and the transplanted organ would likely increase the chance of immediate graft function without DGF. Calcineurin inhibitors (CNI), the mainstay of standard immunosuppression in transplant recipients, are potent vasoconstrictors that may contribute to additional injury leading to long-term development of interstitial fibrosis and tubular atrophy. In this study, the majority of recipients received non-depleting induction and had CNI started early after KT, irrespective of DGF evolution. Delayed initiation of CNI treatment in conjunction with depleting induction therapy may reduce the risk of prolonged DGF (27) (28), and our results trended in this direction. Possibly, novel drug, such as belatacept (29) or eculizumab (30) could significantly contribute to DGF minimization in this context. These therapeutic interventions should be studied prospectively.

Our study was conducted on a population-based sample, allowing generalisation of the results to similar Caucasian populations. The sample size of our study population together with the completeness of the data collected, allowed us to explore new associations between donor and recipient risk factors and DGF. Our study also has some limitations. First, although complete and extensive in terms of parameters evaluated, it represents a retrospective and single centre study.

Second, the definition of DGF (dialysis requirement in the first week after KT) may have biased the results by overestimating DGF events without directly reflecting a marker of ischemic reperfusion injury.

Third, KT from DCD donors to anuric KT recipients clearly increased during the study period (data not shown), so that the groups were not ideally balanced. Therefore, our results are possibly biased by a small percentage of DCD procurement included in our study, and the mid- and long-term outcome results are consequently limited. This is for instance reflected in the very low rate of graft loss in DCD KT to anuric recipients, which is likely the result of selection bias at time of KT.

In conclusion, DCD procurement and recipient residual urine volume are independently associated with DGF rate after KT. These factors are associated with inferior short- and midterm outcome, including increased risk of postoperative complications. DCD procurement and residual diuresis must be included in allocation considerations, not as independent factors, but as an interacting network.

## Data Availability

All data relevant to the study are included in the article or uploaded as supplementary information.

## AUTHOR CONTRIBUTION

FB conducted the literature search, interpreted the results and wrote the manuscript. DS performed the statistical analyses, interpreted the results and thoroughly revised the manuscript. CK, GB, FS and NM participated to conceiving the study. All authors have read and approved this version of the manuscript.

## ETHICS STATEMENT

The study was approved by the Cantonal Ethics Committee of Bern (BASEC-Nr 2020-02754).

## SUPPLEMENTARY MATERIAL

Supplementary Figure 1 and Supplementary Table 1.

