## Supplementary materials for "Impact of marginal donor to marginal recipient kidney transplant on delayed graft function and outcome"

Sppl. Figure 1. Selection procedure. Percentages were calculated using the baseline sample size as denominator. KT: Kidney Transplantation

Total cohort of KT

n = 627

*Exclusion Criteria*

- Living donor KT (n = 208; 33.2%)
- Paediatric recipient (n = 36; 5.7%)
- Multiorgan recipient (n = 16; 2.5%)
- Primary non-function (n = 13; 2%)

Cohort analysed of KT

n = 369 (58.8%)

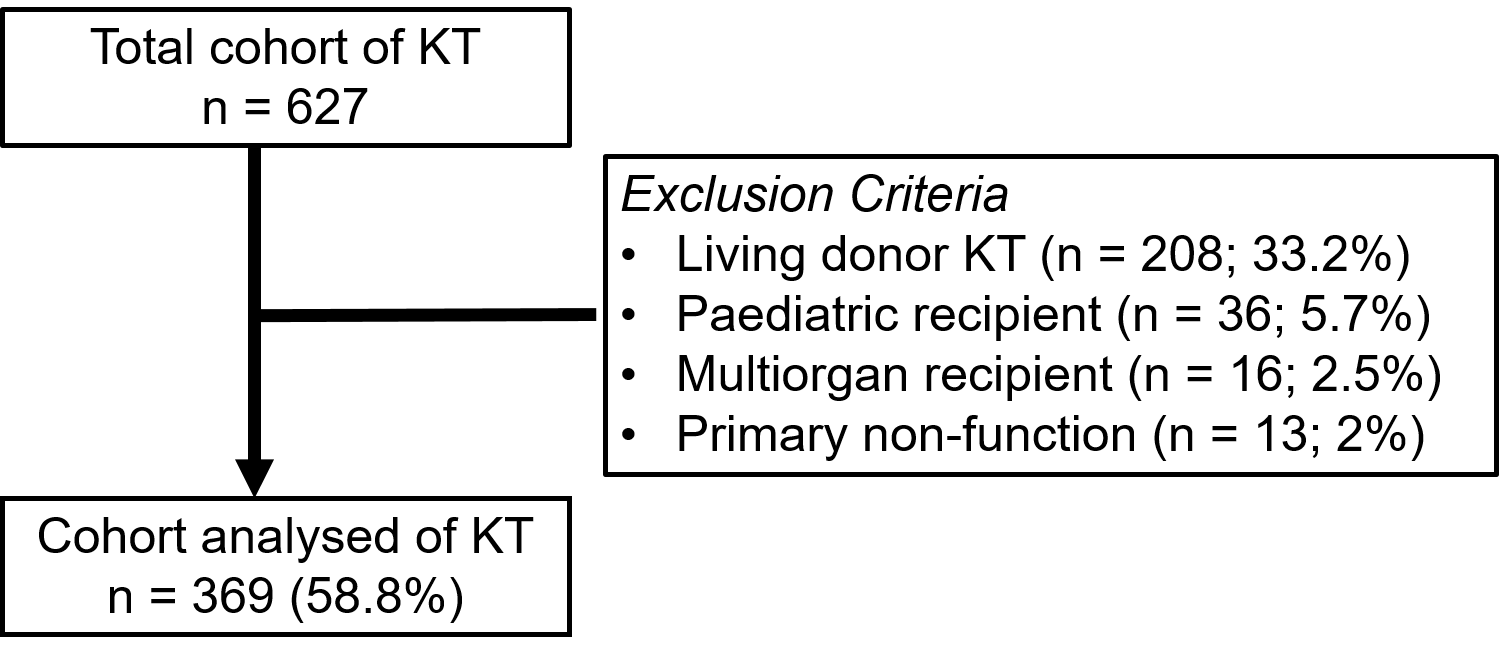

Sppl. Figure 2 Nomogram to predict DGF risk based on donor age, first warm ischemia time, recipient age and residual urine volume. Each factor provides a number points (first row) and the sum of all points is converted to a DGF probability. Nomogram designed from model 2 (Fig 2D-F). DGF: Delayed Graft Function.

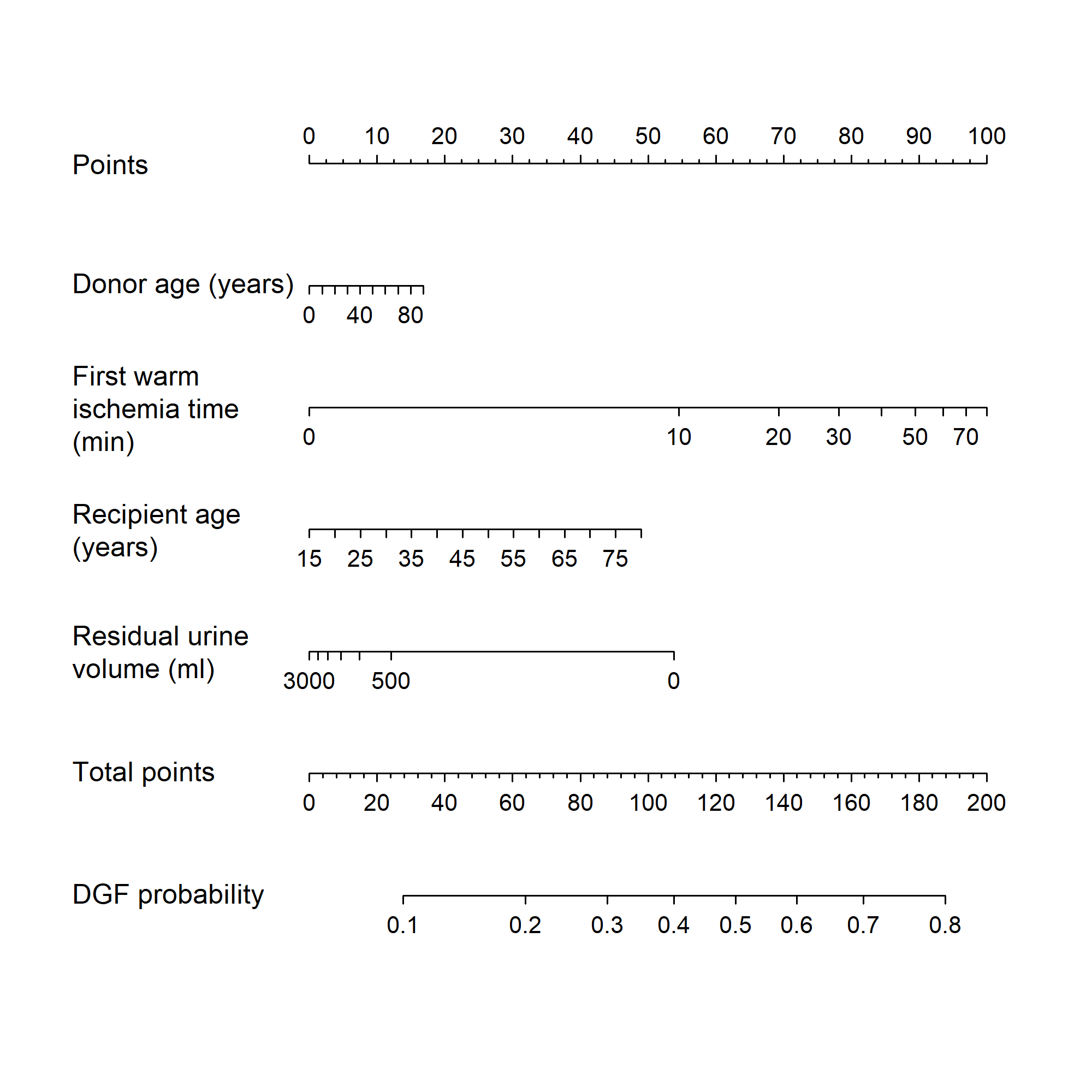

Table 1. Perioperative interventions and DGF risk.

a) Frequency of hypothermic machine perfusion used, depleting induction therapy and pre-KT dialysis sessions for the various groups. KT: Kidney Transplantation; DGF: Delayed Graft Function.

|  | **KT-recipients** | | | | | |
| --- | --- | --- | --- | --- | --- | --- |
|  | Anuria -  DCD -  n = 219 | Anuria - DCD +  n = 39 | Anuria +  DCD -  n = 96 | Anuria +  DCD +  n = 15 | Overall  n = 369 | p-value |
| Hypothermic machine perfusion (yes) | 21 (9.6%) | 19 (49%) | 5 (5.3%) | 6 (40%) | 51 (14%) | **< 0.001** |
| Depleting induction therapy (yes) | 14 (16%) | 8 (26%) | 4 (12%) | 2 (17%) | 28 (18%) | 0.6 |
| Pre-KT dialysis (yes) | 64 (29%) | 14 (36%) | 32 (33%) | 8 (53%) | 118 (32%) | 0.2 |

b) Logistic regression model predicting DGF events from exploratory given variables. DGF: Delayed Graft Function.

|  | OR | 95% CI | p-value |
| --- | --- | --- | --- |
| Hypothermic machine perfusion (yes) | 1.06 | 0.46, 2.33 | 0.9 |
| Depleting induction therapy (yes) | 0.37 | 0.10, 1.12 | 0.10 |
| Pre-KT dialysis (yes) | 1.66 | 0.95, 2.89 | 0.075 |
| Cold ischemia time (per hour) | 1.61 | 0.78, 3.36 | 0.2 |

KT: Kidney Transplantation.
